# The effect of preeclampsia on long-term kidney function among pregnant women with chronic kidney disease

**DOI:** 10.1101/2023.11.16.23298657

**Authors:** Zheng Li, Shi Chen, Ying Tan, Jicheng Lv, Minghui Zhao, Qian Chen, Yingdong He

## Abstract

**Background:** The association between superimposed preeclampsia and an elevated risk of long-term kidney function decline or end-stage renal disease (ESRD) in patients with chronic kidney disease (CKD) remains uncertain. This study aimed to analyze the association between preeclampsia and kidney function deterioration in CKD patients.

**Methods:** This is a retrospective cohort study, included the clinical information of 103 pregnant CKD patients with preeclampsia and 103 matched CKD patients without preeclampsia who were followed-up for a minimum of 1 year after their first pregnancy from January 1, 2009, to May 31, 2022. Cox proportional hazards regression analysis was conducted to evaluate the effects of preeclampsia on long-term kidney function decline or ESRD among CKD patients. Kaplan–Meier curves were used to compare renal survival within different subgroups and compared by the log-rank test.

**Results:** During the follow-up period, 44 (42.72%) CKD patients with preeclampsia and 20 (19.42%) without preeclampsia had an estimated glomerular filtration rate (eGFR) decline >30% or developed ESRD. Compared with CKD patients without preeclampsia, the eGFR declined more significantly in patients with preeclampsia [98.43 (79.48, 116.47) to 81.32 (41.20, 102.97) mL/min/1.73 m^2^ vs. 100.00 (74.86, 120.04) to 89.45 (63.69, 105.60) mL/min/1.73 m^2^; *P*=0.041]. Multivariable analysis showed that early-onset preeclampsia (HR=2.82, 95% CI: 1.48–5.39, *P*<0.01) and late-onset preeclampsia (HR=2.51, 95% CI: 1.28–4.93, *P*<0.05) were both risk factors for an eGFR decline >30% or ESRD.

**Conclusions:** Preeclampsia was associated with a higher risk of long-term kidney function decline or ESRD among CKD patients, especially in patients with early-onset preeclampsia.

**Research in Context:** *Evidence before this study:* Chronic kidney disease (CKD) is proposed as a high-risk factor for preeclampsia, which is an idiopathic disease during pregnancy with multisystemic involvement, including the kidney. It is believed that pregnancy accelerates renal function decline in patients with stage 3-4 CKD. Yet, little is known about whether superimposed PE is associated with an increased risk of renal function decline in patients with CKD. Peking University First Hospital has been paying special attention to the perinatal care of patients with CKD since 2009. Given the high risk of both adverse maternal and neonatal outcomes among women with CKD, multidisciplinary care that includes nephrologists and maternal-fetal medicine specialists was set up in 2018, leading to the referral of more patients with CKD in Beijing and its surrounding areas to our hospital for perinatal care and delivery. Our analysis of the follow-up data of pregnant CKD patients with and without preeclampsia in our hospital over the past 14 years will help us better understand the relationship between preeclampsia and reduction in renal function in patients with CKD.

*Added value of this study:* This longitudinal cohort study including 103 pregnant CKD patients with preeclampsia and 103 matched CKD patients without preeclampsia with minimum follow-up of 1 year, the association between preeclampsia and long-term kidney function decline or ESRD among CKD patients were analyzed. Compared with CKD patients without preeclampsia, the eGFR declined more significantly in patients with preeclampsia [98.43 (79.48, 116.47) to 81.32 (41.20, 102.97) mL/min/1.73 m2 vs. 100.00 (74.86, 120.04) to 89.45 (63.69, 105.60) mL/min/1.73 m2; P=0.041]. Multivariable analysis showed that increased Scr levels (HR=3.02, 95% CI: 1.53–5.94, P=0.001), higher CKD stage (HR=2.76, 95% CI: 1.46–5.22, P=0.002), proteinuria ≥1.00 g/24h (HR=2.70, 95% CI: 1.39–5.25, P=0.003), early-onset preeclampsia (HR=2.82, 95% CI: 1.48–5.39, P<0.01) and late-onset preeclampsia (HR=2.51, 95% CI: 1.28–4.93, P<0.05) were risk factors for an eGFR decline >30% or ESRD.

*Implications of all the available evidence:* This study indicates that preeclampsia was associated with increased risk of eGFR decline>30% or ESRD, especially early-onset preeclampsia. Therefore, for patients with CKD, seeking good prophylactic treatment to prevent the onset of preeclampsia during pregnancy, especially early-onset preeclampsia, is of great significance not only for improving pregnancy outcomes but also for improving long-term prognosis of renal function. The use of LDA to prevent early-onset preeclampsia has a new clinical significance. In addition to improving pregnancy outcomes, LDA may be beneficial for reducing kidney function decline in pregnant CKD patients. This study provided critical evidence to support further prospective studies investigating the association between LDA and long-term kidney function prognosis.

## Introduction

Chronic kidney disease (CKD) is a leading public health problem worldwide, affecting approximately 10% people in the world[1]. Individuals with CKD are at higher risk for preeclampsia. The incidence of preeclampsia is known to be 1-3 times higher in CKD patients (10-15%) than in general population (3-8%)[2]. Thus, to screen and prevent preeclampsia is important for management for pregnant women with CKD.

Several studies have been conducted to explore the long-term effects of pregnancy on renal function in patients with CKD. Pregnancy appeared to accelerate renal function decline in patients with stage 3-4 CKD, however, little effect on renal function decline in patients with stage 1-2 CKD [2–4]. Preeclampsia, which is an idiopathic disease during pregnancy, can lead to systemic multiorgan damage, including hypertension and cardiovascular disease[5, 6]. Whether preeclampsia increases the risk of long-term renal function decline or end stage renal disease (ESRD) remains controversial. Although several studies found that patients superimposed with preeclampsia during pregnancy have an increased risk of ESRD later in life[7, 8], some studies found that preeclampsia does not seem to have an obvious effect on long-term maternal renal function[9–12]. For women with CKD, whether the prognosis of CKD is effect by preeclampsia during pregnancy also remains unclear. The comprehensive exploration of the role of preeclampsia in the risk of renal dysfunction can improve management for pregnant women with CKD.

Here, we have established a cohort of pregnant women with CKD and used propensity score matching based on the presence or absence of preeclampsia to construct our analytical cohort. This study aimed to evaluate the association between preeclampsia and renal function decline or ESRD in patients with CKD.

## Methods

### Study design and participants

The medical information of 703 pregnant women with CKD between January 2009 and May 2022 was collected retrospectively. The inclusion criteria were as follows: (1) preexisting CKD, (2) gestation that proceeded up to the 12th week with complete maternal and infant records, and (3) at least 1 year of follow-up after delivery. The exclusion criteria were patients with pregnancies that ended spontaneously or by therapeutic termination in the first trimester or patients with kidney disease diagnosed during pregnancy. Thirteen patients underwent abortion due to CKD or for personal reasons, 63 patients were transferred to our hospital in the 2^nd^ or 3^rd^ trimester, 16 patients delivered in other hospitals, and 9 patients underwent therapeutic pregnancy termination in the second trimester due to fetal malformation or chromosomal abnormalities. The participants were screened based on the inclusion and exclusion criteria (Figure 1). To evaluate the long-term kidney function outcomes in CKD patients with preeclampsia, we performed a longitudinal study that included 103 CKD patients with preeclampsia and 103 comparable controls. The patients were matched by age, BMI, baseline Scr value, 24-h proteinuria levels and mean arterial pressure (MAP). A total of 206 CKD women with complete pregnancy and childbirth data were enrolled.

**Figure 1.**
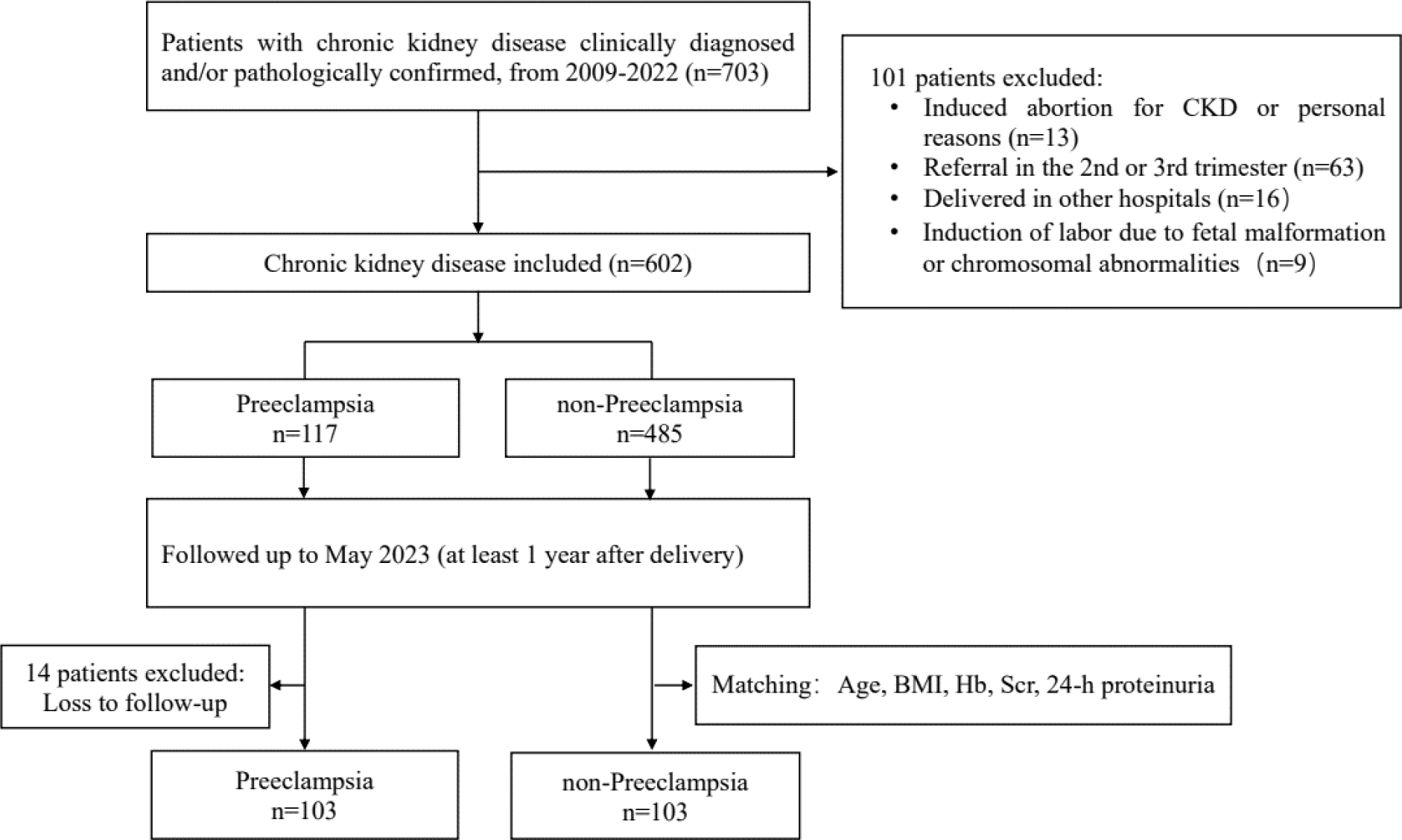
Flow chart of study cohort.

### Ethical approval

The study was undertaken in accordance with the Declaration of Helsinki and approved by the Ethics Committee of Peking University First Hospital [No. 2022 (233)]. The data were anonymous, and the requirement for informed consent was therefore waived.

## Data collection

Clinical and pathological data were extracted for all patients at their first visit to our hospital during pregnancy and follow-up. All patients were followed up every two to four weeks per routine clinical practice during pregnancy and for at least one year postpartum, and clinical information for every visit was obtained from the medical records. The date of the first visit was taken as time point 0, and after delivery, the follow-up time at each visit was calculated (1 to 14 years). Follow-up data were collected through medical record review and outpatient follow-up.

The following baseline information was collected: age, pathological renal biopsy results, CKD type, MAP, body weight, body height, Scr levels, and 24-h proteinuria levels. BMI was calculated as the body weight in kilograms measured during the first exam divided by height in meters squared (kg/m^2^). According to age and Scr levels, the estimated glomerular filtration rate (eGFR) was calculated according to the modification of diet in renal disease (MDRD) equation [13]. For the patients who had undergone renal biopsy, the type of CKD was determined based on the biopsy results; for the other patients, the CKD type was determined based on the clinical diagnosis. The kidney disease stage was classified based on the preconception eGFR as follows: Stage 1: eGFR ≥90.00 mL/(min·1.73 m^2^); Stage 2: eGFR 60.00∼89.00 mL/(min·1.73 m^2^); Stage 3: eGFR 30.00∼59.00 mL/(min·1.73 m^2^); Stage 4: eGFR 15.00∼29.00 mL/(min·1.73 m^2^); and Stage 5: eGFR<15.00 mL/(min·1.73 m^2^) [13].

## Pregnancy outcomes

The adverse pregnancy outcomes included severe preeclampsia, early preterm birth, stillbirth, fetal–neonatal death, very low birth weight infants (VLBWIs) and small for gestational age (SGA). The diagnoses of severe preeclampsia in patients enrolled between 2009 and 2013 were reviewed according to the 2013 guidelines [14].

## Renal outcomes

Kidney disease progression events were defined as a 30% decrease in the eGFR or ESRD without remission after observation for at least 4 weeks or until the end of the follow-up[15, 16]. We also performed multivariable Cox regression analyses based on the outcomes of a 50% decline in eGFR or ESRD. ESRD was defined as an eGFR <15 mL/min/1.73 m^2^ or the initiation of renal replacement therapy (including long-term dialysis or kidney transplantation).

## Definitions

1. CKD was defined as either kidney damage (an albumin-to-creatinine ratio >30.00 mg/24 h for two of three urine specimens, urine sediment abnormalities, tubular disorders, histologically diagnosed abnormalities, structural abnormalities detected by scanning) or a GFR <60.00 mL/min/1.73 m^2^ for three months defined by the Kidney Disease: Improving Global Outcomes (KDIGO) guidelines[13].
2. Preterm birth was defined according to the World Health Organization criteria as all births before 37 completed weeks of gestation. The diagnostic criterion for early preterm birth was a gestational age at birth of less than 34 weeks[17–19].
3. The diagnostic criteria for severe preeclampsia in patients with normal blood pressure and no proteinuria were based on the 2013 Hypertension in Pregnancy Guidelines of the American College of Obstetricians and Gynecologists (ACOG) [14]. For women who had proteinuria but no hypertension in early pregnancy, the diagnosis of severe preeclampsia required the presence of thrombocytopenia, a sudden increase in proteinuria (either five times the baseline value or twice the baseline value if the baseline value exceeded 2.00 g/24 h), hypertension accompanied by severe headaches, epigastric pain, or a serum aspartate aminotransferase concentration greater than 70.00 U/L. For women who had both hypertension and proteinuria in early pregnancy, the diagnosis of severe preeclampsia required any one of the following criteria: an elevated serum aspartate aminotransferase concentration (>70.00 U/L), thrombocytopenia, or worsening hypertension (a systolic blood pressure ≥140.00 mmHg with an increase of at least 30.00 mmHg or a diastolic blood pressure ≥90.00 mmHg with an increase of at least 15.00 mmHg) accompanied by severe headaches or epigastric pain[20, 21].
4. SGA was defined as a birth weight under the 10^th^ percentile based on gestational age[17–19].
5. VLBWIs were defined as neonates with birth weights <1500.00 g [17–19].
6. Stillbirth was defined as the absence of signs of life at or after birth.
7. Neonatal death was defined as death of a live-born neonate during the first 7 days after birth.
8. Controls: To better evaluate the effects of preeclampsia on renal function in patients with CKD, CKD patients without preeclampsia were selected as the control group. The patients were matched by age, BMI, baseline Scr level, 24-h proteinuria level and MAP.

## Statistical analysis

All statistical analyses were conducted with SPSS version 23.0 (IBM, Chicago, IL, USA) and R version 4.2.1 software. Continuous variables are expressed as the mean ± standard deviation (SD) or the median and interquartile range (IQR), and categorical variables are expressed as numbers and percentages. Age, Scr level and MAP were evaluated as dichotomous variables according to the median. Differences in the means between the groups were assessed using the independent samples *t test*, analysis of variance (ANOVA) and Kruskal‒Wallis *H* tests/Wilcoxon’s rank-sum tests for continuous variables, whereas Pearson’s chi-square test or Fisher’s exact test was used for categorical variables. The relevant variables that were significantly associated with progression of renal function decline in the univariable analysis were included in the multivariable models. A multivariable Cox proportional hazard regression analysis was conducted to evaluate the relative risk by generating the hazard ratios (HRs) and 95% confidence intervals (CIs) for preeclampsia and the progression of kidney function decline. We examined the proportional hazards assumption by testing the statistical significance of interactions between the follow-up time and exposures. Kaplan–Meier survival curves were used to compare renal survival within different subgroups and compared by the log-rank test. To better evaluate the effects of preeclampsia on renal function in patients with CKD, the propensity score method was used. Participants were matched using the greedy algorithm with 1:1 pairing. The caliper size was set at 0.025×SD of the logit of the propensity scores. Balanced baseline covariates in the two groups were confirmed by paired comparison tests. A two-sided *P* value <0.05 was considered significant. The Bonferroni correction for multiple comparisons was used.

## Results

### Baseline clinical characteristics

From January 2009 to May 2022, a total of 602 patients with CKD were enrolled in the database. One hundred and seventeen (19.44%) patients developed superimposed preeclampsia, 103 of whom were followed up for at least 1 year. A total of 103 matched patients were selected as the control group. Ultimately, 206 CKD patients were included in this study, with a median maternal age of 32.00 (29.00, 34.00) years at the baseline visit. The median postpartum follow-up time was 5.00 (3.00, 9.00) years. At baseline, 108 (52.43%) CKD patients were in stage 1; 67 (32.52%) were in stage 2; and 31(15.05%) were in stage 3-4. Sixty-three (30.52%) patients had chronic hypertension. Seventy-three (35.44%) patients received low-dose aspirin (LDA) therapy during pregnancy. The baseline clinical information of the two groups was similar. Forty-two (40.78%) patients presented with early-onset preeclampsia and 61 (59.22%) patients presented with late-onset preeclampsia. Table 1 shows the baseline clinical information of the cohort.

**Table 1.**
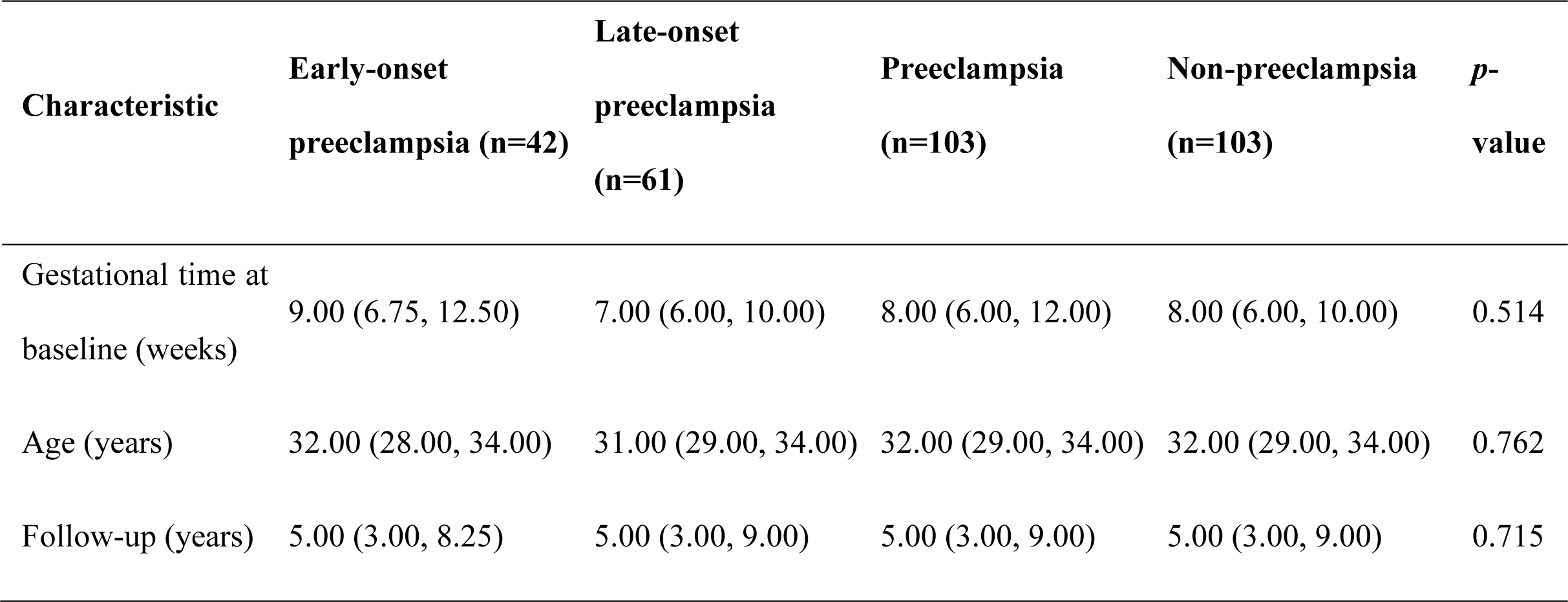
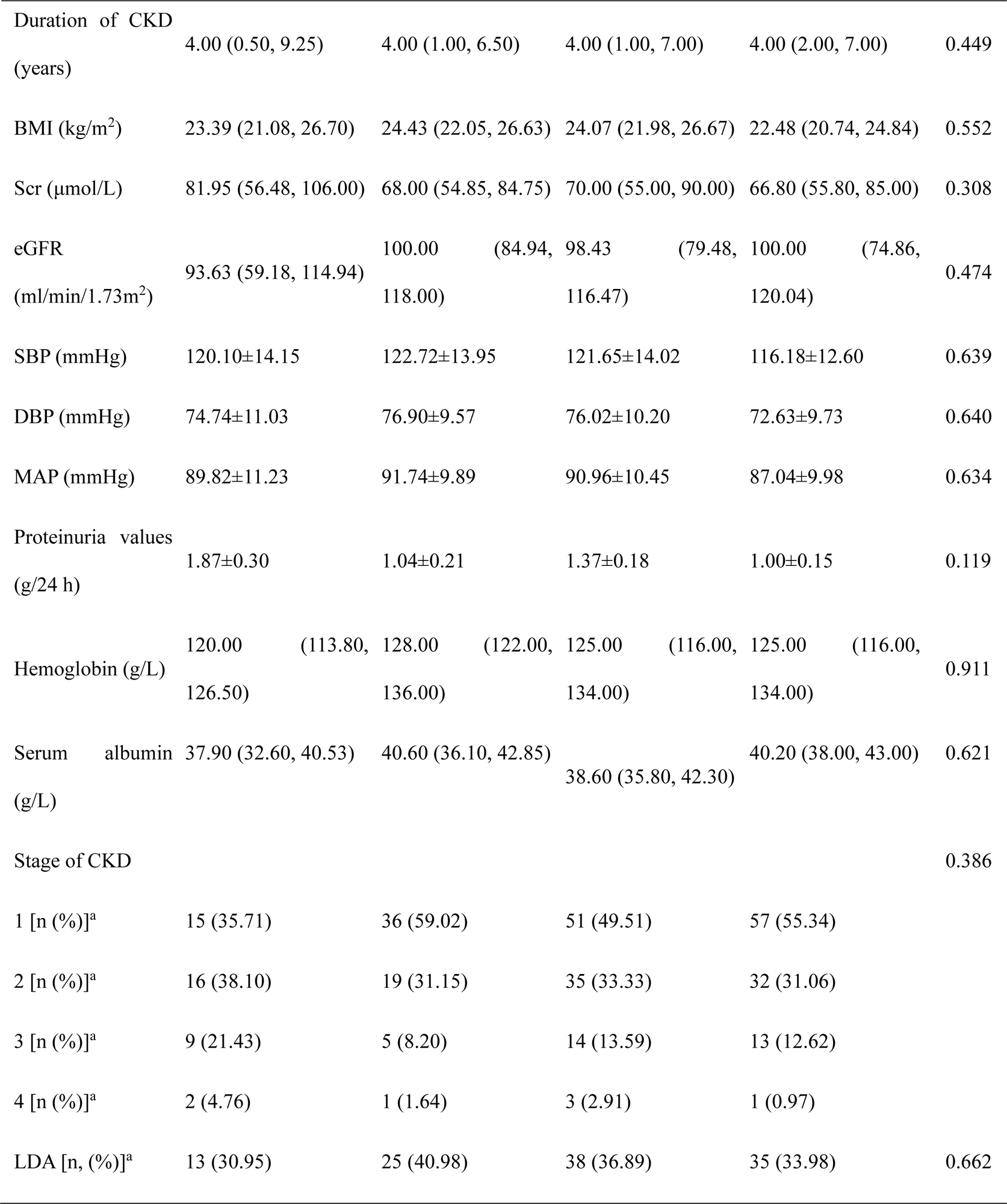
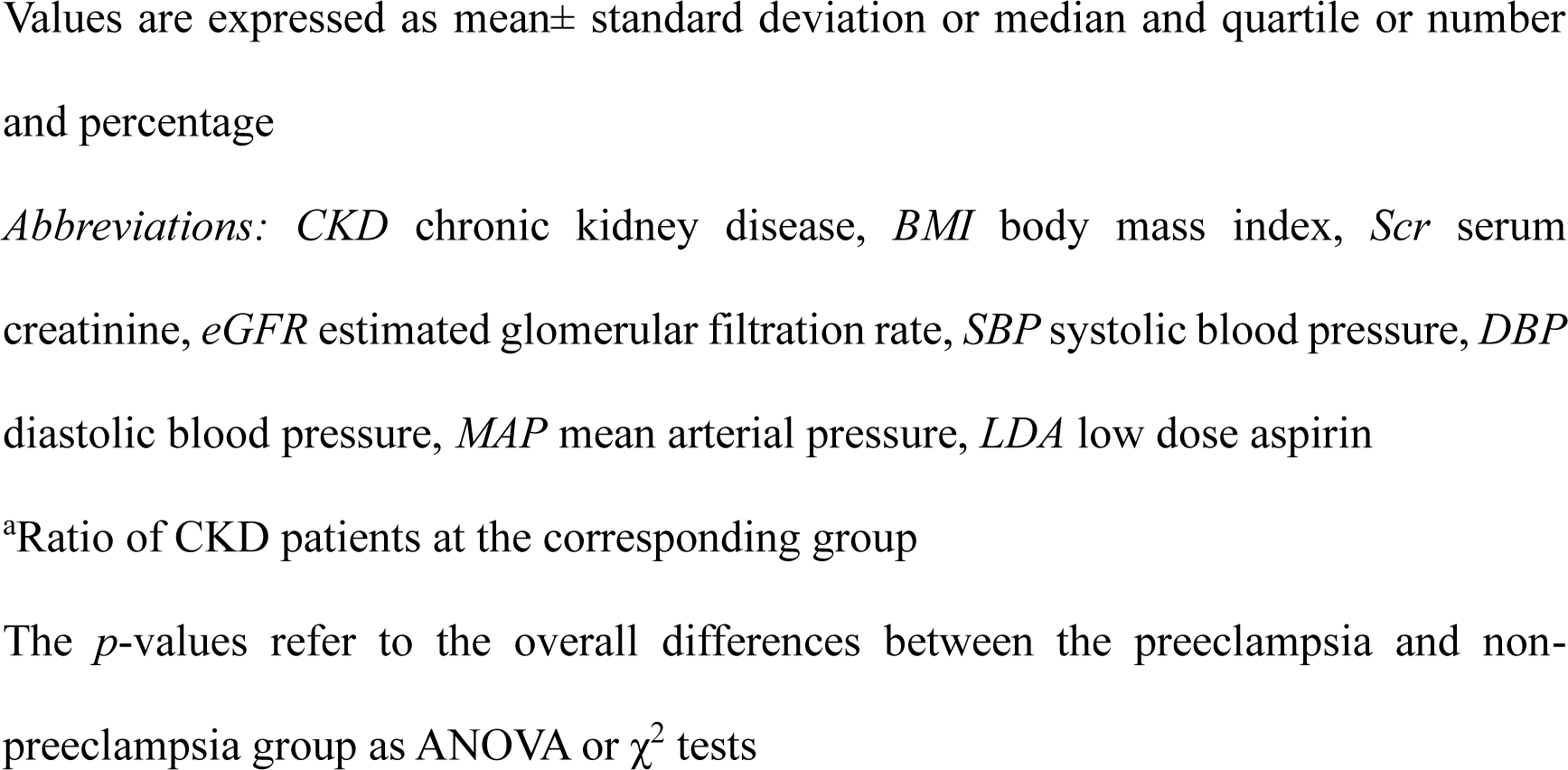
Baseline characteristics of CKD women at first visit during pregnancy.

| **Characteristic** | **Early-onset**<br>**preeclampsia (n=42)** | **Late-onset**<br>**preeclampsia**<br>**(n=61)** | **Preeclampsia**<br>**(n=103)** | **Non-preeclampsia**<br>**(n=103)** | ***p*-**<br>**value** |
| --- | --- | --- | --- | --- | --- |
| Gestational time at<br>baseline (weeks) | 9.00 (6.75, 12.50) | 7.00 (6.00, 10.00) | 8.00 (6.00, 12.00) | 8.00 (6.00, 10.00) | 0.514 |
| Age (years) | 32.00 (28.00, 34.00) | 31.00 (29.00, 34.00) | 32.00 (29.00, 34.00) | 32.00 (29.00, 34.00) | 0.762 |
| Follow-up (years) | 5.00 (3.00, 8.25) | 5.00 (3.00, 9.00) | 5.00 (3.00, 9.00) | 5.00 (3.00, 9.00) | 0.715 |
| Duration of CKD (years) | 4.00 (0.50, 9.25) | 4.00 (1.00, 6.50) | 4.00 (1.00, 7.00) | 4.00 (2.00, 7.00) | 0.449 |
| BMI (kg/m<sup>2</sup>) | 23.39 (21.08, 26.70) | 24.43 (22.05, 26.63) | 24.07 (21.98, 26.67) | 22.48 (20.74, 24.84) | 0.552 |
| Scr (μmol/L) | 81.95 (56.48, 106.00) | 68.00 (54.85, 84.75) | 70.00 (55.00, 90.00) | 66.80 (55.80, 85.00) | 0.308 |
| eGFR (ml/min/1.73m<sup>2</sup>) | 93.63 (59.18, 114.94) | 100.00 (84.94, 118.00) | 98.43 (79.48, 116.47) | 100.00 (74.86, 120.04) | 0.474 |
| SBP (mmHg) | 120.10±14.15 | 122.72±13.95 | 121.65±14.02 | 116.18±12.60 | 0.639 |
| DBP (mmHg) | 74.74±11.03 | 76.90±9.57 | 76.02±10.20 | 72.63±9.73 | 0.640 |
| MAP (mmHg) | 89.82±11.23 | 91.74±9.89 | 90.96±10.45 | 87.04±9.98 | 0.634 |
| Proteinuria values (g/24 h) | 1.87±0.30 | 1.04±0.21 | 1.37±0.18 | 1.00±0.15 | 0.119 |
| Hemoglobin (g/L) | 120.00 (113.80, 126.50) | 128.00 (122.00, 136.00) | 125.00 (116.00, 134.00) | 125.00 (116.00, 134.00) | 0.911 |
| Serum albumin (g/L) | 37.90 (32.60, 40.53) | 40.60 (36.10, 42.85) | 38.60 (35.80, 42.30) | 40.20 (38.00, 43.00) | 0.621 |
| Stage of CKD |  |  |  |  | 0.386 |
| 1 [n (%)]<sup>a</sup> | 15 (35.71) | 36 (59.02) | 51 (49.51) | 57 (55.34) |  |
| 2 [n (%)]<sup>a</sup> | 16 (38.10) | 19 (31.15) | 35 (33.33) | 32 (31.06) |  |
| 3 [n (%)]<sup>a</sup> | 9 (21.43) | 5 (8.20) | 14 (13.59) | 13 (12.62) |  |
| 4 [n (%)]<sup>a</sup> | 2 (4.76) | 1 (1.64) | 3 (2.91) | 1 (0.97) |  |
| LDA [n, (%)]<sup>a</sup> | 13 (30.95) | 25 (40.98) | 38 (36.89) | 35 (33.98) | 0.662 |

Values are expressed as mean ± standard deviation or median and quartile or number and percentage
*Abbreviations:* *CKD* chronic kidney disease, *BMI* body mass index, *Scr* serum creatinine, *eGFR* estimated glomerular filtration rate, *SBP* systolic blood pressure, *DBP* diastolic blood pressure, *MAP* mean arterial pressure, *LDA* low dose aspirin
<sup>a</sup>Ratio of CKD patients at the corresponding group
The *p*-values refer to the overall differences between the preeclampsia and non-preeclampsia group as ANOVA or $\chi^2$ tests

### Adverse pregnancy and renal outcomes

Among pregnant CKD women, the mean gestational age at birth was 35.00 weeks, and the median neonatal birth weight was 2614.93 g. Moreover, compared with CKD patients without preeclampsia, the incidences of perinatal mortality, preterm birth and VLBW were significantly higher among CKD patients with preeclampsia (*P*<0.01). The pregnancy and renal outcomes for the cohort are shown in Table 2 and were stratified according to early-onset and late-onset preeclampsia.

**Table 2.** Pregnancy and renal outcomes of CKD patients with and without preeclampsia.

|  | **Early-onset**<br>**preeclampsia (n=42)** | **Late-onset**<br>**preeclampsia**<br>**(n=61)** | **Preeclampsia**<br>**(n=103)** | **Non-**<br>**preeclampsia**<br>**(n=103)** | ***p*-**<br>**value** |
| --- | --- | --- | --- | --- | --- |
| Twin pregnancy [ n (%) ]<sup>a</sup> | 2 (4.76) | 0 (0) | 2 (1.94) | 2 (1.94) | 0.689 |
| Live birth [ n (%) ]<sup>a</sup> | 36 (83.33) | 61 (100.00) | 97 (94.17) | 101 (98.06) | < 0.01 |
| Live birth infants [n (%) ]<sup>b</sup> | 38 (86.36) | 61 (100.00) | 99 (94.29) | 103 (98.10) | < 0.01 |
| Pregnancy weeks (weeks)<sup>c</sup> | 30.41±2.63 | 37.21±1.37 | 34.39±3.90 | 37.28±3.38 | < 0.01 |
| Mean birth weight (g)<sup>c</sup> | 1360.63±513.16 | 2860.25±477.13 | 2266.34±34.39 | 2960.10±529.73 | < 0.01 |
| Delivery by CS (%)<sup>c</sup> | 36 (85.71) | 43 (70.49) | 79 (81.44) | 62 (60.19) | 0.01 |
| Perinatal death [n (%) ]<sup>a</sup> | 7 (16.67) | 0 (0) | 7 (6.80) | 2 (1.94) | < 0.01 |
| Preterm birth [n, (%) ]<sup>c</sup> | 36 (100) | 17 (27.87) | 53 (54.64) | 21 (20.79) | < 0.01 |
| 28-32 weeks [n (%) ] | 21 (58.33) | 0 (0) | 21 (21.65) | 1 (0.99) | < 0.01 |
| 32-34 weeks [n (%) ] | 12 (33.3) | 0 (0) | 12 (12.37) | 9 (8.91) | < 0.01 |
| LBWI [n, (%) ]<sup>d</sup> | 17 (44.74) | 18 (29.51) | 35 (35.35) | 19 (18.45) | 0.01 |
| VLBWI [n, (%) ]<sup>d</sup> | 22 (57.89) | 0 (0) | 22 (22.22) | 1 (0.97) | < 0.01 |
| SGA [n, (%) ]<sup>d</sup> | 12 (31.58) | 6 (9.84) | 18 (18.18) | 8 (7.77) | 0.01 |
| eGFR decline > 30% or<br>ESRD [ n (%)]<sup>a</sup> | 23 (54.76) | 21 (34.43) | 44 (42.72) | 20 (19.42) | < 0.01 |
| eGFR decline > 50% or<br>ESRD [ n (%)]<sup>a</sup> | 13 (30.95) | 15 (24.59) | 28 (27.18) | 8 (7.77) | < 0.01 |
| ESRD [ n (%)]<sup>a</sup> | 6 (14.29) | 3 (4.92) | 9 (8.74) | 3 (2.91) | 0.044 |

Statistical significance at p=0.005 after Bonferroni correction for multiple comparisons
Values are expressed as mean± standard deviation or median and quartile or number and percentage
*Abbreviations:* *CKD* chronic kidney disease, *LBWI* low birth weight infants, *VLBWI* very low birth weight infants, *SGA* small for gestational age, *eGFR* estimated glomerular filtration rate, *ESRD* end-stage renal disease
<sup>a</sup>Ratio of CKD patients at the corresponding group
<sup>b</sup>Ratio of fetus at the corresponding group
<sup>c</sup>Data were available only in live birth patients
<sup>d</sup>Data were available only in live birth infants
The *p*-values refer to the overall differences between the preeclampsia and non-preeclampsia groups as ANOVA or $\chi^2$ tests

During the follow-up period, forty-four (42.72%) CKD patients with preeclampsia and 20 (19.42%) CKD patients without preeclampsia had an eGFR decline >30% or developed ESRD. Twenty-eight (27.18%) CKD patients with preeclampsia and 8 (17.77%) CKD patients without preeclampsia had an eGFR decline >50% or developed ESRD (Table 2). Overall, in stage 1-2 CKD patients or in stage 3-4 CKD patients, the incidence of progression of renal function decline or ESRD was significantly higher in the preeclampsia group than in the non-preeclampsia group, especially in those with early-onset preeclampsia (*P*<0.01). To better evaluate the effect of preeclampsia on renal function in patients with CKD, the study population was classified into six subgroups: stage 1-2 CKD patients with early-onset preeclampsia, late-onset preeclampsia or without preeclampsia, stage 3-4 CKD patients with early-onset preeclampsia, late-onset preeclampsia or without preeclampsia. Figure 2 shows the incidence of an eGFR decline>30% or ESRD in the different subgroups. Among CKD patients with preeclampsia, one stage 1 CKD patient, three stage 2 CKD patients, two stage 3a CKD patients, two stage 3b CKD patients and one stage 4 CKD patient progressed to ESRD at a median of 5 years after delivery. One of these nine patients underwent kidney transplantation. Two stage 3a CKD patients and one stage 4 CKD patient progressed to ESRD in the group without preeclampsia.

**Figure 2.**
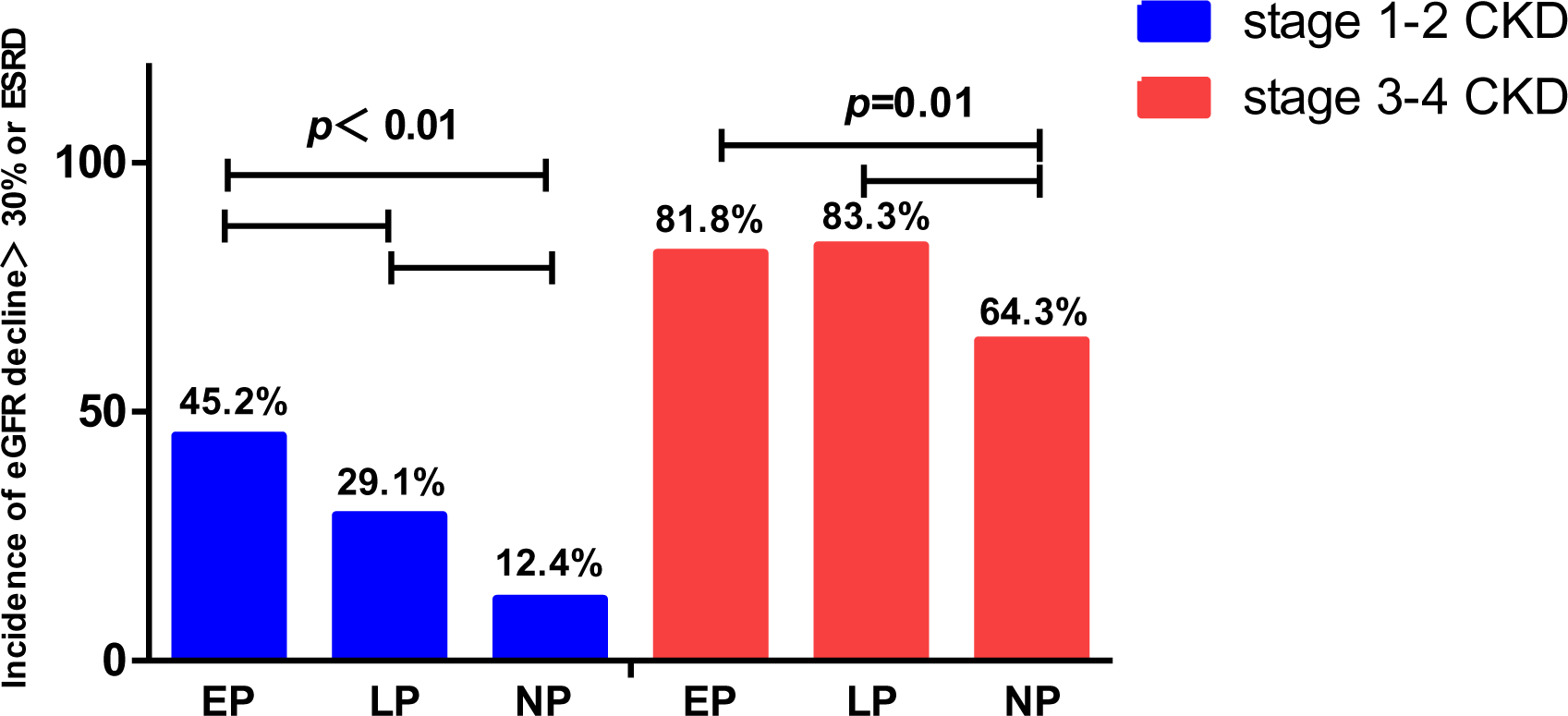
Incidence of eGFR decline <30% or ESRD in different subgroups. During the follow-up period, stage 1-2 CKD patients with preeclampsia (30/86, 34.9%) have a higher risk of eGFR decline>30% or ESRD than stage 1-2 CKD patients without preeclampsia (11/89, 12.4%), *P*> 0.01. Stage 3-4 CKD patients with preeclampsia (14/17, 82.4%) have a higher risk of eGFR decline>30% or ESRD than stage 3-4 CKD patients without preeclampsia (9/14, 64.3%), *P*> 0.01. CKD: chronic kidney disease, eGFR: estimated glomerular filtration rate, ESRD: end stage renal disease, EP: early-onset preeclampsia, LP: late-onset preeclampsia, NP: non-preeclampsia.

### Change in eGFR and Scr level during pregnancy and the follow-up period

During the follow up period, compared with CKD patients without preeclampsia, the eGFR declined more significantly in patients with preeclampsia [98.43 (79.48, 116.47) to 81.32 (41.20, 102.97) mL/min/1.73 m^2^ vs. 100.00 (74.86, 120.04) to 89.45 (63.69, 105.60) mL/min/1.73 m^2^; *P*=0.041]. There was a significant decrease in eGFR [93.63 (59.18, 114.94) to 68.71 (28.62, 90.35) mL/min/1.73 m^2^; *P*<0.05] and an increase in Scr level [81.95 (56.48, 106.00) to 112.95 (74.72, 182.33) μmol/L; *P*<0.05] in CKD patients with early-onset preeclampsia, which was also present in CKD patients with late-onset preeclampsia [eGFR: 100.00 (84.94, 118.00) to 84.06 (47.13, 106.05) mL/min/1.73 m^2^; *P*<0.05], [Scr: 68.00 (54.85, 84.75) to 81.00 (69.00, 117.00) μmol/L; *P*<0.05]. In CKD patients without preeclampsia, we also found a decrease in eGFR [100.00 (74.86, 120.04) to 89.45 (63.69, 105.60) mL/min/1.73 m^2^, *P*<0.05] and an increase in Scr level [66.80 (55.80, 85.00) to 74.00 (63.90, 98.05) μmol/L; *P*<0.05], but less evident than that in CKD patients with preeclampsia. Figure 3 shows the changes in eGFR and Scr level during pregnancy and follow-up period.

**Figure 3.**
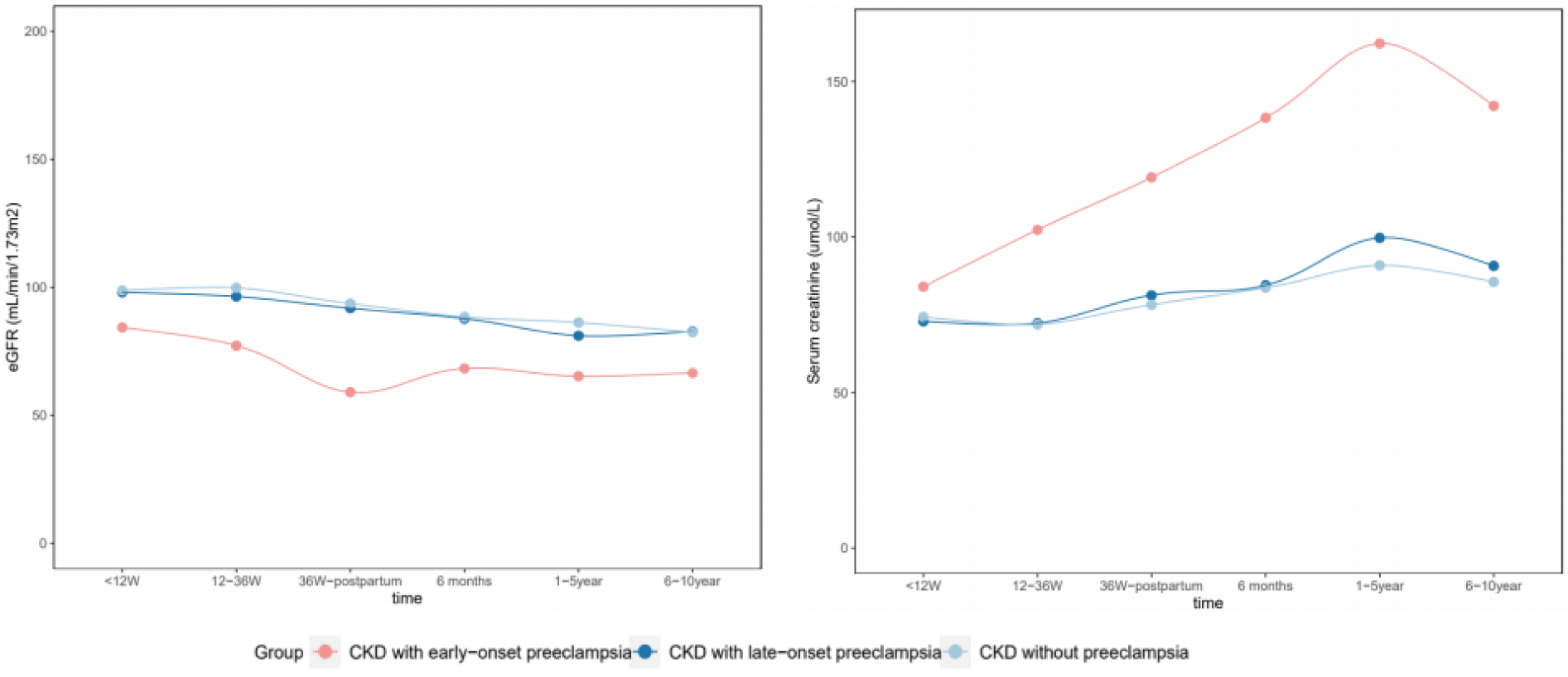
Change in eGFR and serum creatinine in CKD patients with and without preeclampsia. There was a significant decrease in eGFR and an increase in Scr level in CKD women with early-onset preeclampsia (*P*<0.05), which was also present in CKD patients with late-onset preeclampsia(*P*<0.05), but was less evident in CKD patients without preeclampsia.

### Effects of preeclampsia on long-term renal function in patients with CKD

Univariable Cox regression was conducted to assess the relationship between each risk factor and adverse renal outcome, which revealed that hemoglobin levels (HR=0.98, 95% CI: 0.96–0.99, *P*=0.029), serum albumin levels (HR=0.95, 95% CI: 0.91–0.99, *P*=0.016), Scr levels (HR=4.83, 95% CI: 2.68–8.69, *P*<0.01), CKD stage (HR=4.91, 95% CI: 2.90–8.31, *P*<0.01), proteinuria ≥1.00 g/24h (HR=4.22, 95% CI: 2.13–8.35, *P*<0.001), early-onset preeclampsia (HR=3.44, 95% CI: 1.89–6.27, *P*<0.01) and late-onset preeclampsia (HR=1.87, 95% CI: 1.02–3.46, *P*<0.05) were related to an eGFR decline >30% or ESRD. Relevant variables with *P*<0.05 in the univariable analysis and well-recognized risk factors, such as age, BMI, and MAP, were included in the multivariable models even if they were not statistically significant in the univariate analysis. Multivariable analysis showed that increased Scr levels (HR=3.02, 95% CI: 1.53–5.94, *P*=0.001), higher CKD stage (HR=2.76, 95% CI: 1.46–5.22, *P*=0.002), proteinuria ≥1.00 g/24h (HR=2.70, 95% CI: 1.39–5.25, *P*=0.003), early-onset preeclampsia (HR=2.82, 95% CI: 1.48–5.39, *P*<0.01) and late-onset preeclampsia (HR=2.51, 95% CI: 1.28–4.93, *P*<0.05) were risk factors for an eGFR decline >30% or ESRD, as shown in Table 3. Furthermore, increased Scr levels, higher proteinuria level and preeclampsia were associated with an eGFR decline >50% or ESRD, as shown in Table 4. The risk of renal function decline was higher in patients with preeclampsia, especially early-onset preeclampsia, as shown in Figure 4.

**Figure 4.**
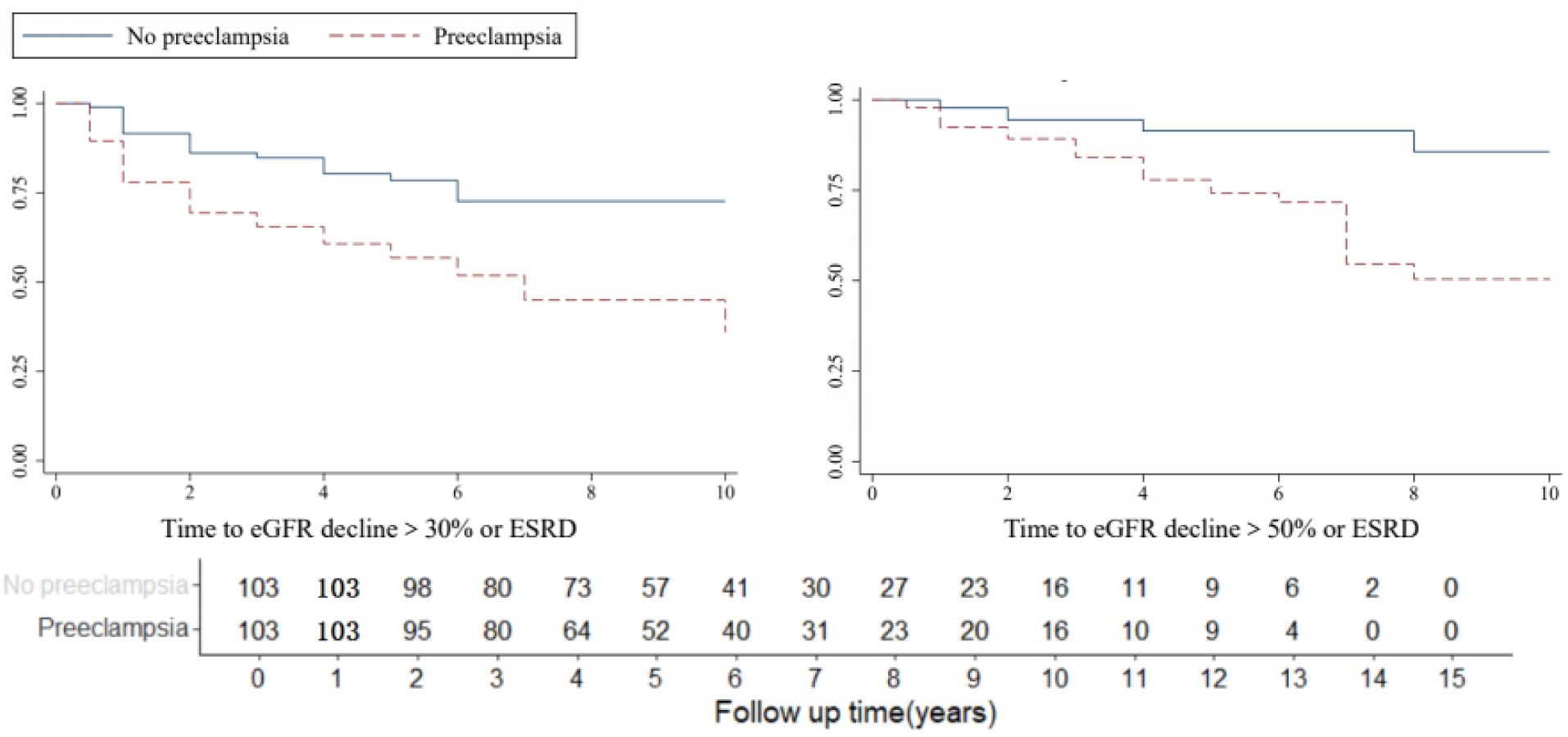
Kaplan-Meier plot showing survival curves for preeclampsia. CKD patients with preeclampsia exhibited a higher risk of eGFR decline>30% or ESRD and eGFR decline>50% or ESRD (*P*<0.05) compared with CKD patients without preeclampsia.

**Table 3.**
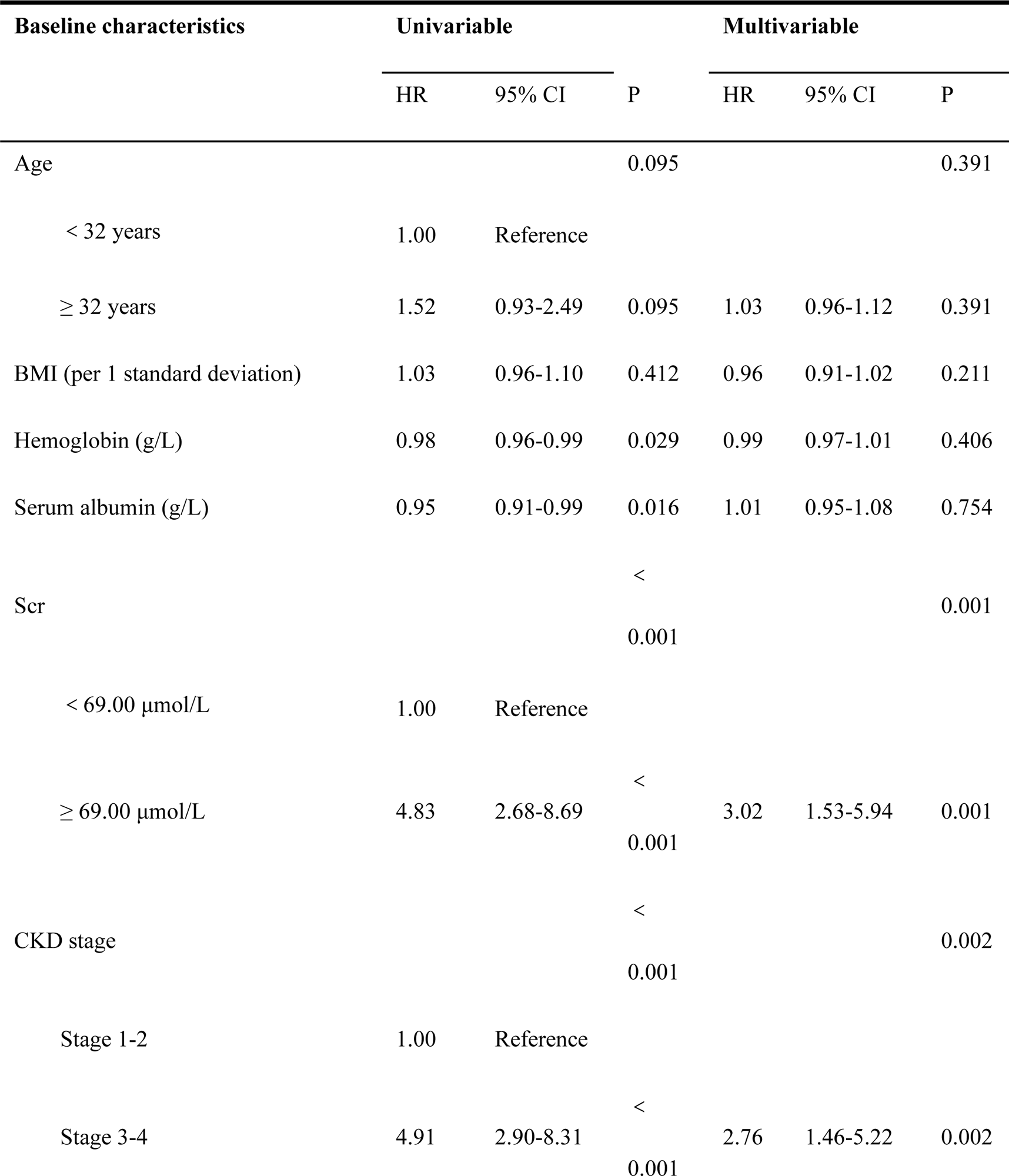
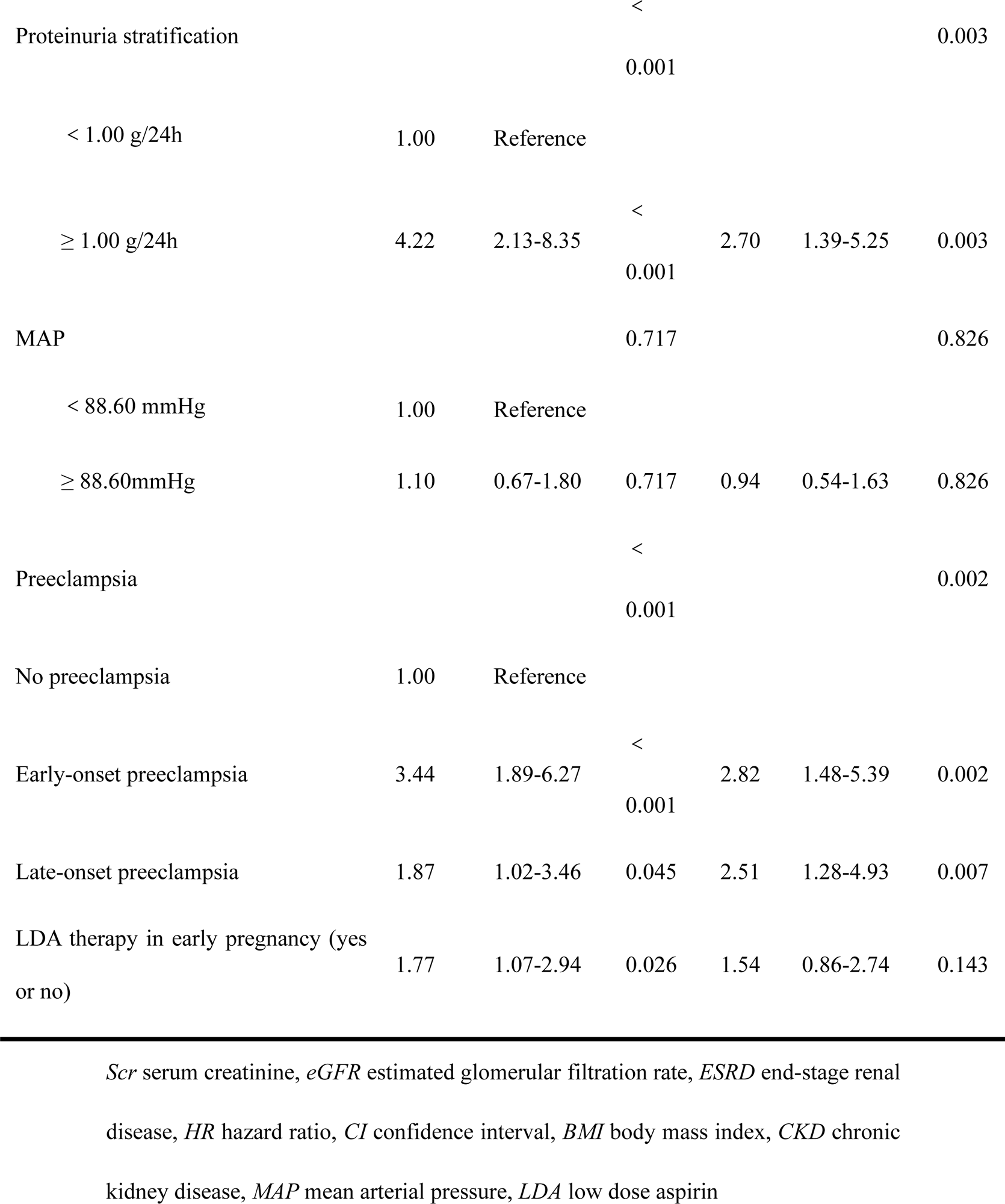
Univariable and multivariable Cox analyses for eGFR decline <30% or ESRD.

**Table 4.** Univariable and multivariable Cox analyses for eGFR decline <50% or ESRD.

| Baseline characteristics | Univariable |  |  | Multivariable |  |  |
| --- | --- | --- | --- | --- | --- | --- |
|  | HR | 95% CI | P | HR | 95% CI | P |
| Age |  |  | 0.99 |  |  | 0.969 |
| < 32 years | 1.00 | Reference |  |  |  |  |
| ≥ 32 years | 1.00 | 0.52-1.94 | 0.99 | 1.00 | 0.91-1.11 | 0.969 |
| BMI (per 1 standard deviation) | 1.05 | 0.97-1.14 | 0.25 | 0.99 | 0.91-1.07 | 0.767 |
| Hemoglobin (g/L) | 9.97 | 0.95-0.99 | 0.004 | 0.98 | 0.96-1.00 | 0.096 |
| Serum albumin (g/L) | 0.91 | 0.86-0.96 | 0.001 | 1.01 | 0.93-1.09 | 0.885 |
| Serum creatinine |  |  | < 0.001 |  |  | 0.027 |
| < 69.00 µmol/L | 1.00 | Reference |  |  |  |  |
| ≥ 69.00 µmol/L | 4.16 | 1.94-8.92 | < 0.001 | 2.70 | 1.12-6.51 | 0.027 |
| CKD stage |  |  | < 0.001 |  |  | 0.118 |
| Stage 1-2 | 1.00 | Reference |  |  |  |  |
| Stage 3-4 | 3.75 | 1.86-7.59 | < 0.001 | 2.09 | 0.83-5.24 | 0.118 |
| Proteinuria stratification |  |  | < 0.001 |  |  | 0.004 |
| < 1.00 g/24h | 1.00 | Reference |  |  |  |  |
| ≥ 1.00 g/24h | 2.73 | 1.66-4.48 | < 0.001 | 3.85 | 1.56-9.53 | 0.004 |
| MAP |  |  | 0.652 |  |  | 0.939 |
| < 88.60 mmHg | 1.00 | Reference |  |  |  |  |
| ≥ 88.60mmHg | 1,17 | 0.60-2.27 | 0.652 | 1.03 | 0.49-2.17 | 0.939 |
| Preeclampsia |  |  | 0.003 |  |  | 0.003 |
| No preeclampsia | 1.00 | Reference |  |  |  |  |
| Early-onset preeclampsia | 4.46 | 1.85-10.77 | 0.001 | 5.07 | 2.00-12.84 | 0.001 |
| Late-onset preeclampsia | 3.21 | 1.36-7.59 | 0.008 | 2.46 | 0.93-6.54 | 0.07 |
| LDA therapy in early pregnancy (yes or no) | 1.43 | 0.71-2.90 | 0.316 | 1.17 | 0.52-2.65 | 0.705 |

*Scr* serum creatinine, *eGFR* estimated glomerular filtration rate, *ESRD* end-stage renal disease, *HR* hazard ratio, *CI* confidence interval, *BMI* body mass index, *CKD* chronic kidney disease, *MAP* mean arterial pressure, *LDA* low dose aspirin

## Discussion

For patients with CKD, pregnancy itself can be a burden on the kidneys. Many previous studies showed that pregnancy could aggravate renal function in patients with CKD, with a higher CKD stage, higher urinary protein levels and chronic hypertension as risk factors for the decline of renal function [17, 22–24]. However, there are few studies on whether superimposed preeclampsia during pregnancy further exacerbates renal function decline in CKD patients. In this study, we found that preeclampsia could accelerate renal function decline in patients with CKD, and this adverse effect was more obvious in patients with stage 3-4 CKD who had worse underlying renal function. Furthermore, we found that the rate of renal function decline was more pronounced in patients with early-onset preeclampsia during pregnancy than in patients with late-onset preeclampsia.

Preeclampsia is an idiopathic disease in pregnancy with the core pathogenesis of systemic vascular endothelial cell activation due to poor immunological regulation at the maternal-fetal interface, which in turn leads to systemic multiorgan damage. Renal damage is one of the specific manifestations of preeclampsia, and increased proteinuria and Scr levels are clinical manifestations for severe preeclampsia. Basic research has shown that most glomerular endotheliosis and podocytopathy caused by preeclampsia is reversible[25–28], and most manifestations related to kidney damage can recover spontaneously after delivery, along with the delivery of the placenta and the recovery of hemodynamic changes. As one of the severe manifestations of hypertensive disorders in pregnancy, superimposed preeclampsia during pregnancy is associated with a significant increased risk of both hypertension and cardiovascular disease in the long term. Some recent population-based studies suggested that women with preeclampsia during pregnancy were at a 5-fold increased risk of ESRD compared with women without preeclampsia[7, 8, 29, 30]. However, there are few studies on whether preeclampsia can further accelerate renal function loss in patients with CKD based on pregnancy status.

Previous studies have found that pregnancy status can accelerate the decline of renal function in women with moderate to severe CKD[31, 32], the worse the underlying renal function is, the faster the decline in renal function. Our study found that, in addition to the burden of pregnancy itself, superimposed preeclampsia will further accelerate the deterioration of renal function, and the accelerated loss of renal function occurs not only in patient with stage 3-4 CKD, but also in stage 1-2 CKD patients. In addition, renal damage in patients with early-onset preeclampsia is more obvious than that in patients with late-onset preeclampsia. Therefore, for patients with CKD, seeking good prophylactic treatment to prevent the onset of preeclampsia during pregnancy, especially early-onset preeclampsia, is of great significance not only for improving pregnancy outcomes but also for improving long-term prognosis of renal function. Currently, LDA is applied to prevent preeclampsia in patients at high risk. A recent study has shown that LDA cannot significantly reduce the incidence of preeclampsia but can delay its onset[33]. In our study, early-onset preeclampsia had a greater effect on renal function decline in patients with CKD than late-onset preeclampsia. Therefore, for pregnant CKD patients, who are high risk population for preeclampsia, the use of LDA to prevent early-onset preeclampsia has a new clinical significance. In addition to improving pregnancy outcomes, LDA may be beneficial for reducing kidney function decline in pregnant CKD patients. However, our previous study found that the use of LDA to prevent preeclampsia in patients with CKD was obvious only in stage 3-4 patients with proteinuria levels greater than 1.00 g[34], suggesting that more precise population screening should be carried out to identify patients with CKD who should receive LDA therapy to prevent preeclampsia. In addition, in this study, the level of proteinuria in the third trimester in CKD patients with preeclampsia was significantly higher than that in CKD patients without preeclampsia. Continuous massive proteinuria is a risk factor for renal function decline. For pregnant patients with superimposed preeclampsia, whether more aggressive control of proteinuria after delivery, rather than waiting for the natural recovery of urine protein as in most obstetric management, can slow renal function decline needs to be further investigated.

There were some limitations of this study. First, this is a retrospective study, which was inevitably affected by sample selection bias. For example, the number of patients with stage 3–4 CKD was small, which limited the study’s power to assess preeclampsia in patients with decreased kidney function[35]. Second, this study did not analyze the effects of different types of kidney disease on pregnancy outcomes and kidney function decline.

## Conclusions

Preeclampsia was associated with a higher risk of long-term kidney function decline or ESRD among CKD patients, especially in patients with early-onset preeclampsia.

## Data Availability

The datasets used and/or analyzed during the current study are available from the corresponding author upon reasonable request.

## Declarations

## Acknowledgments

The authors thank Dr. Yu Sun for her assistance with this study.

## Authorship confirmation statement

All the authors have read the manuscript and approved this submission.

## Ethical approval and statement of patient consent

This retrospective study was approved and monitored by the Institutional Review Board of Peking University Health Science Centre [No. 2022 (233)]. The data were anonymous, and the requirement for informed consent was therefore waived.

## Consent for publication

As this study was a retrospective analysis, formal consent was not required. All the authors of the manuscript agreed to its publication.

## Authors’ contributions

Research idea and study design: ZL, YDH; data acquisition: ZL, YDH; data analysis/interpretation: all authors; statistical analysis: ZL, YDH; supervision or mentorship: JCL, QC and MHZ; funding acquisition: YDH.

## List of abbreviations

ACOG: American College of Obstetricians and Gynecologists
ANOVA: analysis of variance
CIs: confidence intervals
CKD: chronic kidney disease
eGFR: estimated glomerular filtration rate
ESRD: end-stage renal disease
HRs: hazard ratios
IQR: interquartile range
KDIGO: Kidney Disease: Improving Global Outcomes
MAP: mean arterial pressure
MDRD: modification of diet in renal disease
SD: standard deviation
SGA: small for gestational age
VLBWIs: very low birth weight infants

